# Antimycotic Susceptibility of Environmental Airborne Yeasts: Evidence from Diverse Work Environments

**DOI:** 10.1101/2025.06.11.25329417

**Authors:** Adeola Folayan, Stephen Ambu

## Abstract

**Background:** This study compared the susceptibility of environmental yeast isolates from indoor and outdoor air collected at an electronics factory, an office, and a winery in Malaysia to various antifungal agents

**Methods:** Sabouraud’s Dextrose Agar (SDA) (supplemented with 0.05g/L Chloramphenicol to inhibit bacterial growth) and Dichloran Glycerol Agar (DG-18) (for xerophilic yeasts) were used for the isolation of yeasts. Air sample volumes ranging from 10 to 250 litres were drawn using the Ideal Air Sampler (Biomerieux BBL) and impacted onto DG-18 and SDA plates. SDA plates and DG-18 plates were subsequently incubated at 25^°^C and 21^°^C respectively for four days after sampling. Fourteen isolates were randomly selected from each sampling point at each sampling event. Yeast colonies were identified using the commercial biochemical panel Integral Systems Yeast Plus (Liofilchem) after the second subculture from the primary isolation plate. The system also characterises susceptibility to the following antimycotic agents: *nystatin, amphotericin, flucytosine, econazole, ketoconazole, clotrimoxazole, miconazole, itraconazole, voriconazole* and *fluconazole*.

**Results:** A total of 196 yeast isolates from ten species were tested. *Cryptococcus laurentii* was the predominant species across all sites. Most isolates were susceptible to ketoconazole, fluconazole, flucytosine, itraconazole, and nystatin. Lower susceptibility was observed for amphotericin and econazole. Overall, 61.7% of isolates were fully susceptible to all tested agents.

**Conclusions:** The majority of environmental airborne yeasts demonstrated susceptibility to common antimycotics. The dominance of *Cryptococcus laurentii* and the variable susceptibility patterns underscore the need for continued environmental surveillance

## Background

Yeasts have a complex multilayered cell wall structure that confers osmotic protection against lysis. The beta-glucan and chitin are the fibrillar materials in the yeast cell wall. They serve an important function for the rigidity and resistance of the wall. They limit the metabolism of polymers that could cause lysis [1]. The chitin in the yeast cell is also involved in dimorphic transition, especially in *Candida albicans*. Dimorphic transition is implicated in the invasiveness and virulence of yeasts [2]. Yeasts, especially the pathogenic genera like *Candida* are capable of causing various infections such as oral *candidosis*, the most common opportunistic infection in patients with HIV infection and vaginitis, amongst others [3, 4].

An investigation of the responses of yeast isolates to amphotericin B, 5-fluorocytosine and eight azole derivatives showed that all vaginal *Candida albicans* isolates were uniformly sensitive at low concentration to all the antimycotics tested. However, non-*albicans* species, such as *Candida glabrata* and *Saccharomyces cerevisiae*, showed decreased susceptibility to all azoles tested except butoconazole. Other studies have also indicated that the potency of fluconazole and terconazole against non-*albicans in-vitro* is poor [5]. More studies on the responses of emerging pathogenic yeasts such as *Candida glabrata* to commonly prescribed antimycotic agents are on-going [7, 8].

The aim of this study was to compare the sensitivity of different airborne yeast isolates to antimycotic agents. The yeast isolates were obtained from the indoor and outdoor air of different work environments in Malaysia. The environments were an electronic factory, an office and a winery. The winery and the electronic factory were expected to harbor a substantial number of yeast species. Winemaking has been associated with yeast activities, especially *Saccharomyces* spp.. The lubricant used in the selected electronic factory generated an oil mist, which was aerosolized into the air. This oil mist may serve as nutrients for yeasts and other microorganisms, accounting for their abundance indoors in the electronic factory. In view of the abundance of these yeasts indoors, the issue of their antimycotic drug susceptibility is important. This study will also provide valuable data for subsequent studies on the variability in antimycotic drug susceptibility of airborne yeast isolates.

## Methods

Sabouraud’s Dextrose Agar (SDA) (supplemented with 0.05g/L Chloramphenicol to inhibit bacterial growth) and Dichloran Glycerol Agar (DG-18 for xerophilic yeasts) were used for the isolation of yeasts [9, 10]. Yeast sampling was conducted alongside bacterial and fungal sampling, as previously reported in earlier published studies [R]. Six sampling points were located in the electronics factory, five sampling points were located in the winery, and two sampling points were located in the office. There were two outdoor sampling points in the electronics factory. There was just one outdoor sampling point each at the winery and the office. Air sample volumes ranging from 10 to 250 litres were drawn using the Ideal Air Sampler (Biomerieux, BBL). Two sampling events were undertaken between December and May at each site. SDA plates and DG-18 plates were incubated at 25^°^C °C and 21^°^C respectively, for four days after sampling. Fourteen (14) isolates were randomly selected from each sampling point at each sampling event. Wet mounts were prepared to identify yeast colonies before proceeding with identification. Yeast colonies were identified with Integral Systems Yeast Plus, ISYP (Liofilchem), using the second subculture from the primary isolation plate. Other than identification, ISYP also characterises susceptibility to the antimycotic agents, nystatin, amphotericin, flucytosine, econazole, ketoconazole, clotrimoxazole, miconazole, itraconazole, voriconazole and fluconazole.

## Results

A total of 196 yeast cultures isolated from the indoor and outdoor air of three different work environments were investigated to determine their sensitivity to 11 antimycotic agents. Yeast isolates were most sensitive to *ketoconazole* followed by *fluconazole, flucytosine, itraconazole*, and *nystatin*, and least sensitive to *amphotericin and econazole* (Tables 1 and 2). The isolates represented 10 species. *Candida* spp. represented 6.3% whilst *Cryptococcus* spp. represented 93.7% of the total yeasts isolated inside the electronic factory. Twenty percent (20%) of the isolates outside the electronic factory were *Candida* spp. whilst the remaining 80% of the isolates were *Cryptococcus* spp. All the isolates identified inside the office were *Cryptococcus* spp.. *Trichosporon pullulans* represented 3.7%, *Candida* spp. 7.4% and *Cryptococcus* spp. 88.9% inside the winery. *Cryptococcus laurentii* was the dominant species at the three study sites, both indoors and outdoors. At least 50% of the isolates at each sampling site, both indoors and outdoors, were susceptible to all the antimycotic agents studied. Sixty-one percent (61.7%) of the isolates from this study were susceptible to the 11 antimycotic agents tested. No isolate was totally resistant to all the antimycotic agents but some isolates were resistant to one or more antimycotics tested.

**Table 1:** Proportion and percentages of environmental yeast isolates susceptible to antimycotic agents.

|  | Isolates susceptible to all eleven antimycotic agents |  | Isolates with intermediate resistance to one antimycotic agent |  | Isolates resistant to one antimycotic agent |  | Isolates resistant to two or more antimycotic agents |  |
| --- | --- | --- | --- | --- | --- | --- | --- | --- |
|  | Proportion | Percentage (%) | Proportion | Percentage (%) | Proportion | Percentage (%) | Proportion | Percentage (%) |
| **Yeast isolates from the indoor of the electronic factory** |  |  |  |  |  |  |  |  |
| All Species | 86/144 | 59.7 | 39/144 | 27.1 | 24/144 | 16.7 | 7/144 | 4.8 |
| *Candida famata* | 0/2 | 0 | 1/2 | 50 | 1/2 | 50 | 0/2 | 0 |
| *Candida glabrata* | 1/1 | 100 | 0/1 | 0 | 0/1 | 0 | 0/1 | 0 |
| *Candida lambica* | 1/1 | 100 | 0/1 | 0 | 0/1 | 0 | 0/1 | 0 |
| *Candida parapsilosis* | 3/3 | 100 | 0/3 | 0 | 0/3 | 0 | 0/3 | 0 |
| *Candida rugosa* | 2/2 | 100 | 0/2 | 0 | 0/2 | 0 | 0/2 | 0 |
| *Cryptococcus albidus* | 8/12 | 66.7 | 2/12 | 25 | 3/12 | 37.7 | 2/12 | 25 |
| *Cryptococcus laurentii* | 68/118 | 57.6 | 34/118 | 28.8 | 19/118 | 16.1 | 5/118 | 4.2 |
| *Cryptococcus neoformans* | 1/1 | 100 | 0/1 | 0 | 0/1 | 0 | 0/1 | 0 |
| *Cryptococcus uniguttulatus* | 2/4 | 50 | 2/4 | 50 | 1/4 | 25 | 0/4 | 0 |
| **Yeast isolates from outdoors of the electronic factory** |  |  |  |  |  |  |  |  |
| All species | 8/10 | 80 | 1/10 | 10 | 1/10 | 10 | 0/10 | 0 |
| *Candida lambica* | 1/1 | 100 | 0/1 | 0 | 0/1 | 0 | 0/1 | 0 |
| *Candida rugosa* | 1/1 | 100 | 0/1 | 0 | 0/1 | 0 | 0/1 | 0 |
| *Cryptococcus albidus* | 2/2 | 100 | 0/2 | 0 | 0/2 | 0 | 0/2 | 0 |
| *Cryptococcus laurentii* | 4/6 | 66.7 | 1/6 | 16.7 | 1/6 | 16.7 | 0/6 | 0 |
| **Yeast isolates from indoors of the office** |  |  |  |  |  |  |  |  |
| *Cryptococcus laurentii* | 4/8 | 50 | 3/8 | 37.5 | 1/8 | 12.5 | 0/8 | 0 |
| **Yeast isolates from indoors of the winery** |  |  |  |  |  |  |  |  |
| All specie | 24/27 | 88.9 | 3/27 | 11.1 | 0/27 | 0 | 0/27 | 0 |
| *Candida famata* | 2/2 | 100 | 0/2 | 0 | 0/2 | 0 | 0/2 | 0 |
| *Trichosporon pullulans* | 1/1 | 100 | 0/1 | 0 | 0/1 | 0 | 0/1 | 0 |
| *Cryptococcus albidus* | 3/3 | 100 | 0/3 | 0 | 0/3 | 0 | 0/3 | 0 |
| *Cryptococcus laurentii* | 18/21 | 85.7 | 3/21 | 14.3 | 0/21 | 0 | 0/21 | 0 |
| **Yeast isolates from indoors of the winery** |  |  |  |  |  |  |  |  |
| All specie | 6/7 | 85.7 | 1/7 | 14.2 | 0/7 | 0 | 0/7 | 0 |
| *Candida famata* | 2/2 | 100 | 0/2 | 0 | 0/2 | 0 | 0/2 | 0 |
| *Cryptococcus laurentii* | 4/5 | 80 | 1/5 | 20 | 0/5 | 0 | 0/5 | 0 |

Note: No yeast was isolated from the office outdoors

**Table 2:**
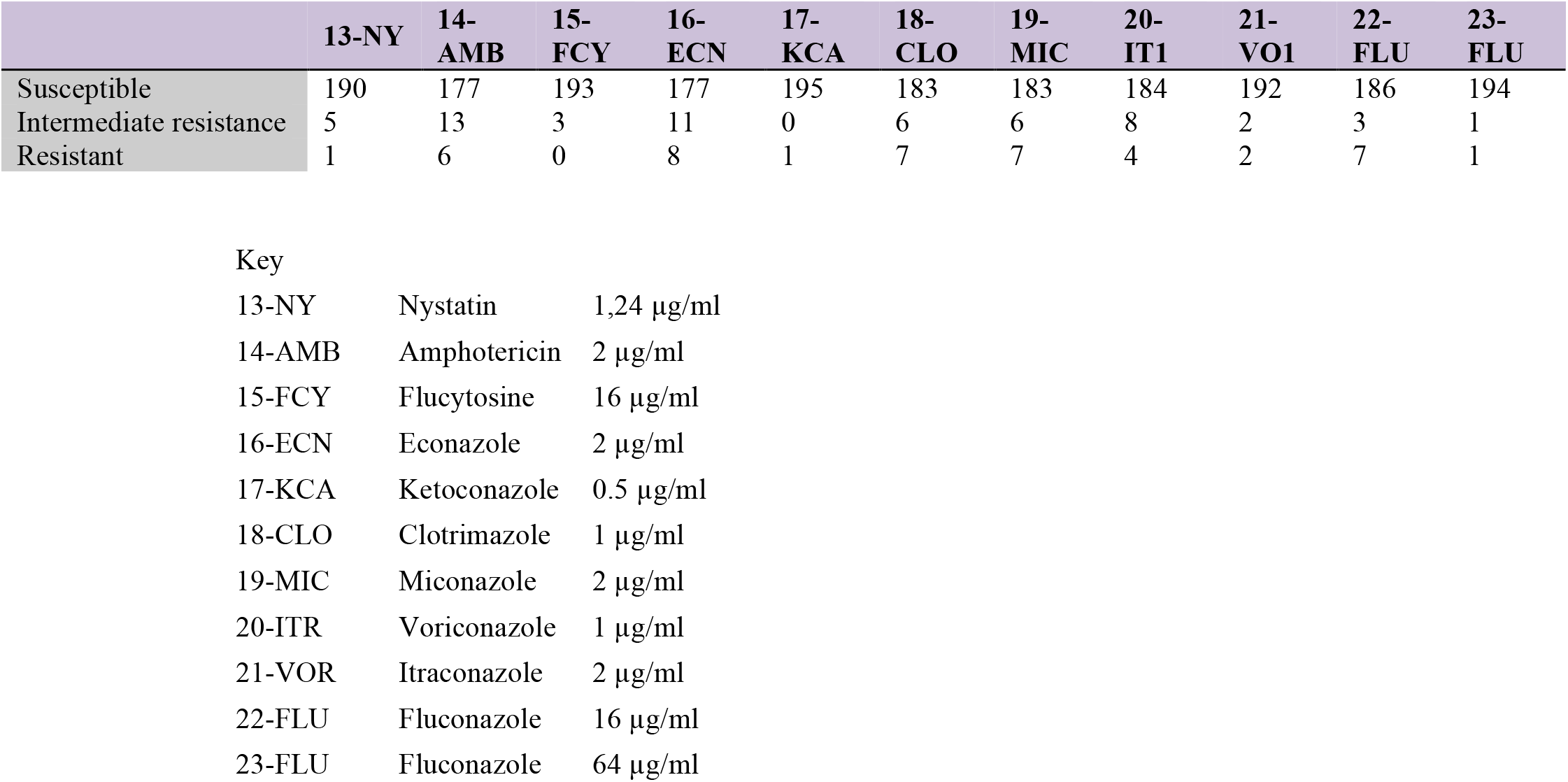
Number of isolates susceptible, intermediately resistant, and resistant to antimycotics of the 196 isolates investigated.

|  | **13-NY** | **14-AMB** | **15-FCY** | **16-ECN** | **17-KCA** | **18-CLO** | **19-MIC** | **20-IT1** | **21-VO1** | **22-FLU** | **23-FLU** |
| --- | --- | --- | --- | --- | --- | --- | --- | --- | --- | --- | --- |
| Susceptible | 190 | 177 | 193 | 177 | 195 | 183 | 183 | 184 | 192 | 186 | 194 |
| Intermediate resistance | 5 | 13 | 3 | 11 | 0 | 6 | 6 | 8 | 2 | 3 | 1 |
| Resistant | 1 | 6 | 0 | 8 | 1 | 7 | 7 | 4 | 2 | 7 | 1 |

Key
|  |  |  |
| --- | --- | --- |
| 13-NY | Nystatin | 1,24 µg/ml |
| 14-AMB | Amphotericin | 2 µg/ml |
| 15-FCY | Flucytosine | 16 µg/ml |
| 16-ECN | Econazole | 2 µg/ml |
| 17-KCA | Ketoconazole | 0.5 µg/ml |
| 18-CLO | Clotrimazole | 1 µg/ml |
| 19-MIC | Miconazole | 2 µg/ml |
| 20-ITR | Voriconazole | 1 µg/ml |
| 21-VOR | Itraconazole | 2 µg/ml |
| 22-FLU | Fluconazole | 16 µg/ml |
| 23-FLU | Fluconazole | 64 µg/ml |

## Discussion

Our findings align with previous studies indicating the variable susceptibility of environmental yeasts to antimycotics [13, 14). Medeiros *et al* examined isolates from tropical freshwater in Brazil. They reported that yeast isolates susceptibility of 13% to ketoconazole, 79% to fluconazole, and 31% to terbinafine [13] Milanezi *et al*., in their study on the susceptibility of yeast isolates from water samples to antifungals in Porto Alegre, found that 16.8% of the yeast isolates were resistant to amphotericin B. They isolated potentially pathogenic species with antifungal resistance and those with promising biotechnological applications. Their study highlights the importance of assessing biological diversity in degraded environments and examining the relationship between contaminants and yeast adaptation [14]. *Cryptococcus laurentii* was the dominant species in this study, in line with the study by Leite *et al*. They reported *Cryptococcus spp* isolates as the dominant from the dust microhabitat in Brazilian libraries [15].

Although the susceptibility of isolates varied in this study, most isolates were susceptible to antimycotics; however, the presence of partially resistant isolates might suggest a potential environmental reservoir for resistant yeasts. Continuous monitoring of airborne yeasts and their susceptibility profiles is crucial for safeguarding public health. There are limited studies to date on the susceptibility of environmental airborne yeasts, especially in work and industrial environments. This highlights the need for further research to better understand their diversity, antifungal resistance patterns, and potential health risks in occupational settings.

## Conclusion

This study highlights the importance of environmental surveillance for airborne yeasts, particularly in diverse work environments. Despite variability in susceptibility among the isolates, the predominance of susceptible strains was evident; however, the identification of partially resistant isolates raises concerns about a potential environmental reservoir for antifungal-resistant yeasts.

## List of abbreviations used

SDA: Sabouraud’s dextrose agar
DG-18: Dichloran glycerol agar

## Authors’ contributions

AF undertook the literature review, performed the bench work and wrote the first draft of this paper. SA advised on the study design and points of discussion and contributed their broad expertise in the field of study. All authors read and approved the final manuscript.

## Availability of data

All data produced in the present study are available upon reasonable request to the authors

## Acknowledgements

We would like to thank the International Medical University (IMU), Malaysia, for their financial support for this research.

## References

1. Ene IV, Walker LA, Schiavone M, Lee KK, Martin-Yken H, Dague E, Gow NA, Munro CA, Brown AJ. Cell wall remodeling enzymes modulate fungal cell wall elasticity and osmotic stress resistance. MBio. 2015 Sep 1;6(4):10–128.

2. Cassone A: Cell wall of pathogenic yeasts and implications for antimycotic therapy. Drugs Exp Clin Res. 1986; 12(6-7): 635–43

3. Holmstrup P, Axéll T: Classification and clinical manifestations of oral yeast infections. Acta Odontol Scand 1990.48(1): 57–59

4. Mrudula P, Joanne TS, Maeve MC: Effect of antifungal treatment on the prevalence of yeasts in HIV-infected subjects. J Med Microbiol 2006.55:1279–128

5. Lynch M E, Sobel J D. Comparative in vitro activity of antimycotic agents against pathogenic vaginal yeast isolates. Med Mycol 1994.32(4): 267–274

6. Canilo-Mufloz A J, Tur C, Torres J: In-vitro antifungal activity of sertaconazole, bifonazole, ketoconazole and miconazole against yeasts of the Candida genus. J Antimicrob Chemoth 1996.37: 815–819

7. Buzzini P, Corazzi L, Turchetti B, Buratta M, Martini A: Characterization of the in vitro antimycotic activity of a novel killer protein from Williopsissaturnus DBVPG 4561 against emerging pathogenic yeasts. FEMS Microbiol Lett 2004; 238: 359–365

8. Turchetti B, Pinelli P, Buzzini P, Romani A, Heimler D, Franconi F, Martini A: In vitroantimycotic activity of some plant extracts towards yeast and yeast-like strains. Phytother Res. 2005.19(1): 44–49

9. Cooley JD, Wong WC, Jumper CA, Straus DC: Correlation between the prevalence of certain fungi and sick building syndrome.Occup. Environ. Med. 1998; 55: 579–584

10. Radon K, Danuser B, Iversen M, Monso E, Weber C, Hartung J, Donham K, Palmgren U, Nowak D: Air contaminants in different European farming environments. Ann Agric Environ Med 2002; 9(1): 41–48

11. Folayan A, Mohandas K, Ambu S, Kumarasamy V, Lee N, Mak JW. Kytococcus sedentarius and Micrococcus luteus: highly prevalent in indoor air and potentially deadly to the immunocompromised–should standards be set?. Tropical Biomedicine. 2018;35(1):149–60.

12. Folayan A, Ambu S. Indoor air bacterial and fungi bioburden in an electronic factory, an office and a winery in Malaysia. Chulalongkorn Medical Journal. 2020;64(2):211–9.

13. Medeiros AO, Kohler LM, Hamdan JS, Missagia BS, Barbosa FA, Rosa CA. Diversity and antifungal susceptibility of yeasts from tropical freshwater environments in Southeastern Brazil. Water Research. 2008 Aug 1;42(14):3921–9.

14. Milanezi AC, Witusk JP, van der Sand ST. Antifungal susceptibility of yeasts isolated from anthropogenic watershed. Anais da Academia Brasileira de Ciências. 2018 Dec 17;91(1):e20170369.

15. Leite DP, Amadio JV, Martins ER, Simões SA, Yamamoto AC, Leal-Santos FA, Takahara DT, Hahn RC. Cryptococcus spp isolated from dust microhabitat in Brazilian libraries. Journal of Occupational Medicine and Toxicology. 2012 Dec;7:1–7.

16. Ferreira-Paim K, Andrade-Silva L, Mora DJ, Lages-Silva E, Pedrosa AL, Da Silva PR, Andrade AA, Silva-Vergara ML. Antifungal susceptibility, enzymatic activity, PCR-fingerprinting and ITS sequencing of environmental Cryptococcus laurentii isolates from Uberaba, Minas Gerais, Brazil. Mycopathologia. 2012 Jul;174:41–52.

